# Predictors of bovine *Schistosoma japonicum* infection in rural Sichuan, China

**DOI:** 10.1101/2021.08.20.21262368

**Authors:** Elise Grover, Sara Paull, Katerina Kechris, Andrea Buchwald, Katherine James, Yang Liu, Elizabeth J. Carlton

## Abstract

In China, bovines are believed to be the most common animal source of human schistosomiasis infections, though little is known about what factors promote bovine infections. The current body of literature features inconsistent, and sometimes contradictory results, and to date, few studies have looked beyond physical characteristics to identify the broader environmental conditions that predict bovine schistosomiasis. Because schistosomiasis is a sanitation-related, water-borne disease transmitted by many animals, we hypothesized that several environmental factors − such as the lack of improved sanitation systems, or participation in agricultural production that is water-intensive − could promote schistosomiasis infection in bovines. Using data collected as a part of a repeat cross-sectional study conducted in rural villages in Sichuan, China from 2007 to 2016, we used a Random Forests, machine learning approach to identify the best physical and environmental predictors of bovine *S. japonicum* infection. Candidate predictors included: 1) physical/biological characteristics of bovines, 2) human sources of environmental schistosomes, 3) socio-economic indicators, 4) animal reservoirs, and 5) agricultural practices. The density of bovines in a village and agricultural practices such as the area of rice and dry summer crops planted and the use of night soil as an agricultural fertilizer, were among the top predictors of bovine *S. japonicum* infection in all collection years. Additionally, human infection prevalence, pig ownership and bovine age were found to be strong predictors of bovine infection in at least one year. Our findings highlight that presumptively treating bovines in villages with high bovine density or human infection prevalence may help to interrupt transmission. Furthermore, village-level predictors were stronger predictors of bovine infection than household-level predictors, suggesting future investigations may need to apply a broad ecological lens to identify potential underlying sources of persistent transmission.

## 1. Introduction

Bovines have long-been recognized as a key contributor to human schistosomiasis in Asia (Gray et al., 2007; Guo et al., 2006), and emerging evidence suggests they may be contributing to human schistosomiasis in Africa (Colley and Loker, 2018).Schistosomiasis is among the most burdensome helminth infections worldwide, with transmission being reported in a total of 78 countries in 2018 and approximately 230 million people in need of preventative treatment (World Health Organization, 2020). Although great strides have been made in the last few decades in global schistosomiasis control (World Health Organization, 2020), pockets of reemergent or persistent transmission highlight the need for careful consideration of possible local drivers of transmission (Kittur et al., 2019; Song et al., 2016). A poignant example is found in *Schistosoma japonicum* transmission in China, where despite well-established control programs and considerable progress towards elimination since the mid-1950s (Xu et al., 2016), a national report highlighted that there remained 450 endemic counties in 2020, a quarter of which had not achieved elimination criteria by year’s end (Zhang et al., 2021). *S. japonicum* is transmitted by at least 40 species of wild and domestic mammals (Gray et al., 2009b), and animal activities near likely transmission sites may be important sources of reemergence and persistence. Although there are several domesticated and wild animals that can carry and transmit *S. japonicum*, (Li et al., 2015; Van Dorssen et al., 2017), estimates from Eastern China suggest that bovines may be responsible for as much as 75% of human transmission (Guo et al., 2006). This substantial contribution is thought to be related to the high degree of environmental overlap between humans and bovines during agricultural production, as well as the large amount of fecal output of bovines, which is estimated to be as high as 100 times that of human fecal production each day (Gray et al., 2009b; He et al., 2001; Ross et al., 2001). Additionally, the high frequency of livestock movement via the livestock trade within mountainous regions of China may facilitate the spread of *S. japonicum* infections across the region (Zheng et al., 2000; Zhou et al., 2012).

Bovine parasitic flatworm infections also have important veterinary impacts. The consequences of *Schistosoma bovis* and other parasitic flatworms can be quite severe on the animal, including: mortality, extreme morbidity and organ damage, emaciation, reduced milk yields and other production issues, and greater susceptibility to other pathogens (Al-Gaabary et al., 2009; Alemneh et al., 2015; McCauley et al., 1983).

Despite strong evidence that bovines are an important driver of human *S. japonicum* infection (Colley and Loker, 2018; Gray et al., 2007; Guo et al., 2006; Rudge et al., 2013) and of human schistosomiasis in Africa, there are major gaps in our understanding of the factors that contribute to bovine *Schistosoma* infection. Most studies assessing bovine infection risks have focused primarily on veterinary schistosome species, the results of which have been inconsistent. For example, several recent assessments of *Schistosoma bovis* infection risk in Eastern Africa assessed bovine sex, age, breed and body condition as predictors of bovine infection status with contradictory results (Chanie et al., 2012; Defersha and Belete, 2018; Gebremeskel et al., 2017; Kebede et al., 2018; Lulie and Guadu, 2014; Tsega and Derso, 2015; Yihunie et al., 2019). Whereas assessments from 2012 and 2017 in Northwest Ethiopia found that neither age, sex nor the bovine’s body condition was associated with bovine schistosomiasis infection (Chanie et al., 2012; Gebremeskel et al., 2017), a 2018 assessment found no significant effect of sex or breed, but suggested that bovines with poor body condition and bovines aged 2-5 had the highest infection risk (Defersha and Belete, 2018). By contrast, a 2019 study from northwestern Ethiopia recently highlighted that female sex, breed and poor body condition were all associated with bovine infection, while age was not found to be associated (Yihunie et al., 2019). Reasons for such discrepancies have not been fully elucidated, though Defersha & Belete (2018) hypothesize that it may be related to variations in management practices for different bovine groups (e.g. separation of sexes or of age groups) and different grazing ranges and patterns allowed on different farms (e.g. smaller grazing area of very young and very old bovines) (Defersha and Belete, 2018).

Outside of eastern Africa, few studies have characterized predictors of bovine schistosomiasis infection. One study from Malaysia found that older age, low weight and male sex, were all risk factors for *Schistosoma spindale* infection in a range of different cattle species (Tan et al., 2015). By comparison, a study conducted primarily among water buffaloes (96.2% water buffalos, 3.8% cattle) in Southern China found that *S. japonicum* infection intensity was highest in young bovines (<2 years) (Gray et al., 2007). These seemingly contradictory results may potentially be explained by isolation and limited grazing for calves in Malaysia (Tan et al., 2015), as well as potential genus-related differences in acquired immunity and self-cure rates (Li et al., 2014), as studies assessing worm establishment success in the two genera have found that cattle are more susceptible to infection than water buffalo (He et al., 1992; Xu et al., 1993). Nevertheless, water buffaloes may still act as important hosts in marshland areas of China, as they are more likely to spend time in water, and therefore more likely to be involved in the *S. japonicum* transmission cycle (He et al., 2001).

Not only is there a great deal of disagreement in the current body of literature over the key risk factors for bovine schistosomiasis infection, but studies to date have almost exclusively focused on physical characteristics rather than broader environmental conditions. There is a considerable literature that documents the role of social and environmental conditions in human schistosomiasis. For example, water, sanitation and hygiene (WASH) infrastructure, agriculture and fishing production, irrigation and night soil use that − is, the collection of either treated or untreated human and/or animal waste for use as fertilizer − have all been implicated as significant risk factors for human *Schistosoma* infection (Carlton et al., 2015; Grimes et al., 2015; Southgate, 1997; Zhou et al., 2018). While it is plausible that many of these factors pose similar risks to bovines, this has not yet been tested. Given that bovine *parasitic flatworm* infections can yield significant morbidity and economic losses, and contribute substantially to human *S. japonicum* infection risk, studies aimed at assessing potential predictors of *S. japonicum* infection in bovines are of paramount importance.

To address this gap, we set out to assess a range of potential physical and environmental predictors of bovine *S. japonicum* infection in 2007, 2010 and 2016 at the individual, household and village-levels in a region where schistosomiasis persistence has been demonstrated to exist in both humans and bovine populations. We draw from the literature on human and bovine schistosomiasis to define a broad set of candidate predictors that describe individual bovine characteristics. In so doing, our study helps to fill the gaps in knowledge about the conditions that predict *S. japonicum* infection in bovines, as well as the relevance of two additional assessment scales (household and village-level) that have rarely been considered in the studies conducted to date. The results of this study will ultimately serve as a critical stepping stone toward developing appropriately-scaled and targeted schistosomiasis intervention and control activities.

## 2. Materials and Methods

### 2.1 Village selection

This study was conducted in two rural counties in Sichuan, China where schistosomiasis reemerged and persisted despite aggressive control efforts. Surveys of environmental and social risk factors, as well as human and bovine infection were conducted in 2007, 2010 and 2016. In 2007, three of eight Sichuan counties where schistosomiasis had reemerged (Liang et al., 2006), were selected for inclusion in our study based on surveillance record availability and the local control stations’ willingness and capacity to collaborate, and a set of 17 to 19 villages was selected in each county, as described previously (Carlton et al., 2011). However, in May 2008, a 7.9 magnitude earthquake severely impacted one of the study counties (Carlton et al., 2015), such that, in 2010, follow-up surveys were conducted in 2 counties (36 villages). In an effort to hone in on the highest risk locations within the two study counties in a region where infections were declining, surveillance records were reexamined in 2016, from which, a total of ten villages were selected for inclusion in the 2016 data collection. Of these, seven were villages that were also included in 2007 and 2010, while three were new. We restrict this analysis to the two counties surveyed in all three years, noting that our villages are not a representative sample of Sichuan, but rather a survey of high-risk locations where schistosomiasis has persisted despite control efforts. All villages are located in the hilly regions of rural Sichuan and ranged from approximately 20 to 150 households and 50 to 200 residents. Finally, the analysis presented here was restricted to villages where bovines were both present and tested for *S. japonicum* infection: 35 villages in 2007, 30 villages in 2010 and 8 villages in 2016. Details about the total number of included villages, households and bovines are provided in Supplementary Table S1. *2.2 Census, household questionnaires and infection surveys*

A village census was conducted in each collection year and all residents over the age of five were invited to participate in surveys for *S. japonicum* infection. In addition, attempts were made to survey all bovines in the village for *S. japonicum* infection. In the summers of 2007, 2010 and 2016, the head of each household was asked to complete a household survey that contained closed-ended questions related to socioeconomic status, domestic and farm animal ownership, sanitation and water access and agricultural practices. Bovine age, type and sex were collected at the time of the bovine infection surveys in 2007 and 2010 (these data were not collected in 2016). Trained staff from the Sichuan Center for Disease Control and Prevention and the county Schistosomiasis Control Stations piloted and conducted all surveys in the local Sichuan dialect. Only bovines that had both household survey data and *S. japonicum* infection data were included in this assessment (Supplementary Table S1).

*S. japonicum* infection surveys were conducted by attempting to test three stool samples on three consecutive days from eligible humans and all bovines in the village. Infection surveys were conducted in November and December of 2007 and 2010, and July 2016. Bovines were isolated in a pen or tied up until a stool was produced on three separate days (consecutive, when possible). All stool samples were transported to the central laboratory soon after collection to be examined using the miracidium hatching test, following standard protocols (Department of Disease Control, 2000). To account for the short survival and rapid hatching of bovine miracidia, the bovine samples were examined for miracidia at one, three and five hours after preparation for at least two minutes each time, whereas human samples were assessed at two, five and eight hours after preparation. One sample from each human was also examined using the Kato Katz thick smear procedure in 2007 and 2010 (Katz et al., 1972). A bovine was considered positive for *S. japonicum* if any miracidium hatching test was positive. A human was considered positive for *S. japonicum* if any miracidium hatching test or the Kato-Katz test was positive. Each person who tested positive was notified and referred to the local anti-schistosomiasis control station for treatment. When bovines tested positive, owners were notified and the animal was referred to the county veterinary station for treatment.

### 2.3 Ethics statement

This study was approved by the Sichuan Institutional Review Board, the University of California, Berkeley, Committee for the Protection of Human Subjects, and the Colorado Multiple Institutional Review Board. All participants provided written, informed consent. The collection of bovine samples was determined to be exempt from review by the Animal Care and Use Committee at the University of California, Berkeley and the Institutional Animal Care and Use Committee at the University of Colorado Anschutz.

### 2.4 Predictor selection and definitions

The primary outcome of interest in this analysis was bovine *S. japonicum* infection in 2007, 2010 and 2016. All candidate predictors were defined using either the household surveys or the human infection surveys and were divided into five categories: 1) biological/physical characteristics; 2) potential human sources of environmental schistosomes; 3) socio-economic indicators; 4) potential animal reservoirs/sources of infection; 5) agricultural risk factors (Table 1). We included agriculture as its own category because bovines are frequently employed in agricultural work in China (Zheng et al., 2000), and because different crop types and agricultural practices have their own inherent exposure risks (e.g. planting wet crops like rice may increase the likelihood of contact with snail habitat and exposure to cercariae (Gordon et al., 2019)). Variables with hypothesized similar mechanisms of transmission risk were aggregated where possible. Three crop type categories were created: winter crops (primarily rapeseed and wheat), summer dry crops (primarily corn, peanuts and vegetables) and summer wet crops (rice). Night soil use was also included as an agricultural risk factor and divided into three categories: night soil use on winter crops, dry summer crops and wet summer crops.

**Table 1.** Summary of predictor variables included in the analysis.

| Predictor list | Predictor category <sup>a</sup> | Scale of analysis | Years available | Variable type |
| --- | --- | --- | --- | --- |
| Bovine type (water buffalo vs. cow) | Physical/biological | Individual | 2007, 2010 | Binary |
| Bovine sex | Physical/biological | Individual | 2007, 2010 | Binary |
| Bovine age | Physical/biological | Individual | 2007, 2010 | Continuous |
| Number of hatch tests <sup>b</sup> | Physical/biological | Individual | All | Discrete |
| County of residence | Physical/biological | Household | All | Binary |
| Prevalence of human infection in village | Potential human sources | Village | All | Continuous |
| Number of infected human household members | Potential human sources | Household | All | Discrete |
| Percent of households with improved sanitation | Socio-economic indicators | Village | All | Continuous |
| Household has improved sanitation (y/n) <sup>c</sup> | Socio-economic indicators | Household | All | Binary |
| Village mean asset score (0-9) | Socio-economic indicators | Village | All | Continuous |
| Household asset score (0-9) | Socio-economic indicators | Household | All | Discrete |
| Percent of households that own cats | Animal reservoirs/sources | Village | All | Continuous |
| Household cat ownership | Animal reservoirs/sources | Household | All | Binary |
| Percent of households that own dogs | Animal reservoirs/sources | Village | All | Continuous |
| Household dog ownership | Animal reservoirs/sources | Household | 2007, 2010 | Binary |
| Mean number of pigs per household | Animal reservoirs/sources | Village | 2010, 2016 | Continuous |
| Household pig ownership | Animal reservoirs/sources | Household | 2010 | Discrete |
| Mean number of bovines per household | Animal reservoirs/sources | Village | All | Continuous |
| Household owns other bovines | Animal reservoirs/sources | Household | All | Discrete |
| Mean area of rice planted per household | Agricultural risk factors | Village | All | Continuous |
| Area of rice planted by household | Agricultural risk factors | Household | All | Continuous |
| Mean area of dry summer crop planted per household | Agricultural risk factors | Village | All | Continuous |
| Area of dry summer crop planted by household | Agricultural risk factors | Household | All | Continuous |
| Mean area of winter crop planted per household | Agricultural risk factors | Village | All | Continuous |
| Area of winter crop planted by household | Agricultural risk factors | Household | All | Continuous |
| Mean number of buckets of night soil used on rice crops | Agricultural risk factors | Village | All | Continuous |
| Number of buckets of night soil used on rice crops | Agricultural risk factors | Household | All | Discrete |
| Mean number of buckets of night soil used on dry summer crops | Agricultural risk factors | Village | All | Continuous |
| Number of buckets of night soil used on dry summer crops | Agricultural risk factors | Household | All | Discrete |
| Mean number of buckets of night soil used on winter crops | Agricultural risk factors | Village | All | Continuous |
| Number of buckets of night soil used on winter crops | Agricultural risk factors | Household | All | Discrete |

<sup>a</sup> Predictors were grouped into five categories relevant to bovine *S. japonicum* infection risk probability: 1) physical/biological characteristics (e.g. old-age); 2) potential human sources of environmental schistosomes (e.g. human *S. japonicum* infection prevalence in the bovine's household); 3) socio-economic indicators (e.g. prevalence of improved sanitation systems in the surround village); 4) potential animal reservoirs/sources of infections (e.g. prevalence of dog ownership in the surrounding village); 5) agricultural risk factors (e.g. a household's total rice crop area).
<sup>b</sup> Because not all bovines produced three stool samples, and examination of a greater number of stool samples can increase the probability of detecting infection, the number of hatch tests used on a given bovine was also included as a predictor.
<sup>c</sup> Improved sanitation was defined as access to an improved toilet in the household, including a biogas digester or a three-compartment toilet.

There were minor variations in the household survey content and question formulation across the study period (e.g., pig ownership was not assessed in 2007). Where possible, continuous/discrete predictors were included over binary measures. We excluded binary variables from the analysis if they represented very rare (<10%) or very common conditions (>90%). Continuous variables were excluded when >90% of the observations took a single value. For example, household dog and pig ownership were both excluded in 2016 because >90% of the households owned dogs while >90% did not own any pigs. A composite household asset score (0-9) was developed for use in this assessment, which included eight household assets assessed in all three collection years (washing machine, television, air conditioner, refrigerator, computer, car, motorcycle), as well as a binary measure indicating that the home was constructed of concrete, wood or bricks (vs. adobe).

Because prior work has demonstrated that group-level measures can serve as important predictors of schistosomiasis infection in humans (Carlton et al., 2015), we also generated village-level predictors from the household survey data. Village-level variables represent all households that participated in the household survey from a given village, even if they didn’t own bovines. Village-level variables were either the village-average value of continuous household measures, or for binary variables, the proportion of the village population reporting the condition. Notably, the village-level variables excluded all observations from the bovine’s own household, and instead used only the data from the other households in the village that participated in the household survey. This allowed for an assessment of how the surrounding village environment impacts individual bovine infection risk, independent of the home environment, whereas the household-level variables aim to unpack the influence of the unique household environment on bovine infection status. The aforementioned criteria led to a total of 31 predictors.

### 2.5 Analysis

Among the bovines with *S. japonicum* infection data (i.e. the outcome variable), those that lacked all household survey data (i.e. the predictor variables) were excluded from the analysis (30/503 bovines in 2007; 36/233 bovines in 2010; and 1/72 in 2016). Infection prevalence was similar among the excluded bovines (11/67, 16.4%) as compared to those included in this analysis (111/741, 15.0%). Among included bovines, missingness was generally low: the variable with the most missing data was bovine sex in 2007 (21.6% missing). Missing values were imputed separately for each collection year using the rflmpute function from the “randomForest” package in R (Breiman, 2018; RColorBrewer and Liaw).

Two preliminary descriptive analyses were carried out. First, spatial patterns of bovine infection prevalence by village were visualized using ESRI’s ArcGIS ArcMap software release 10.5.1 (Environmental Systems Research Institute (ESRI), 2017). Second, categorical versions of the candidate predictors were generated in order to evaluate changes in the distribution of predictors over time as well as to allow a qualitative examination of the distribution of predictors by bovine infection status.

To determine which of our candidate predictors serve as the best predictors of bovine schistosomiasis infection in 2007, 2010 and 2016, a random forests (RF) machine learning approach was used. For each year, 25% of the data was reserved for validation, while the remaining 75% was used for model construction. To address class imbalance in our outcome of interest (13.3%, 17.3%, and 19.7% of bovines were *S. japonicum* positive in 2007, 2010 and 2016, respectively), over sampling of the minority class was conducted. For model tuning, 10-fold cross validation was performed using the Caret package in R to help select the optimal maximum node size and the number of variables to try at each branch. Once the optimal value of each of these parameters was determined, a final model was run using 5000 trees per forest (Breiman and Cutler, 2011).

For each collection year, we conducted a total of ten rebalancing and model tuning iterations to assess the degree of stability in our variable importance rankings. The mean decrease in accuracy (MDA) value was used to rank the top ten predictors from each model on a scale of ten to one − from most to least important − such that the predictor with the highest MDA was assigned a score of 10, and predictors not in the top ten were assigned a score of zero. These variable rankings were then summed across the 10 rebalancing iterations to give a 10-model summary score of variable importance, ranging from 100-1. The ten highest scoring variables from the ten-model summary score were then reassigned a final ranking of first to tenth. Next, we created a “lean” ranking, using only those predictors ranked first through tenth within each collection year. To do this, we performed an additional ten iterations of the aforementioned balancing and tuning process with the top ten predictors, thereby reducing excess noise in the variable ranking assessment caused by including a large number of candidate predictors. Because we hypothesized that the inclusion of human infection as a predictor of bovine infection would strongly influence the predictive capacity of our RF models due to a presumed association between bovine and human infection, we also conducted ten iterations of a sensitivity analysis for each collection year that excluded the human infection variables from the algorithm. The ability of the full, lean and sensitivity RF models to predict infection status was assessed using accuracy, sensitivity and the receiver operator curve (ROC) area under the curve (AUC), respectively. In the case of disagreement or a tie when comparing the chosen performance metrics, kappa and specificity were subsequently compared to select the top performing model for each year.

Simple logistic regression analyses were performed to assess the direction of association between the top predictors and bovine infection, dividing continuous variables into tertiles to assess for potential non-linear relationships. The direction of association was recorded for the top predictors within each collection year, using a p-value of <0.2 to indicate weak evidence of a between-group difference. In the case that no difference was indicated between tertile groups at the p<0.2 level, the predictors were further divided into quartiles and re-assessed. If still no evidence for a between group difference was identified using quartiles, this point was noted in the results. Density plots by infection status were also examined for a subset of predictors that were found to have a change in the direction of association across the collection years. Stata 15 and R Studio 4.0 were used for all analyses (RStudio Team, 2020; StataCorp, 2015).

## 3. Results

### 3.1 Bovine infection prevalence and population characteristics

Villages with high bovine infection prevalence (>30%) were present in all collection years. While some villages were found to have repeatedly high infection prevalence in bovines across all years despite treatment programs and aggressive control measures, other villages emerged as high prevalence villages in 2016 after years of low infection prevalence in bovines. Figure 1 shows a map of bovine infection distribution by village across two counties.

**Figure 1.**
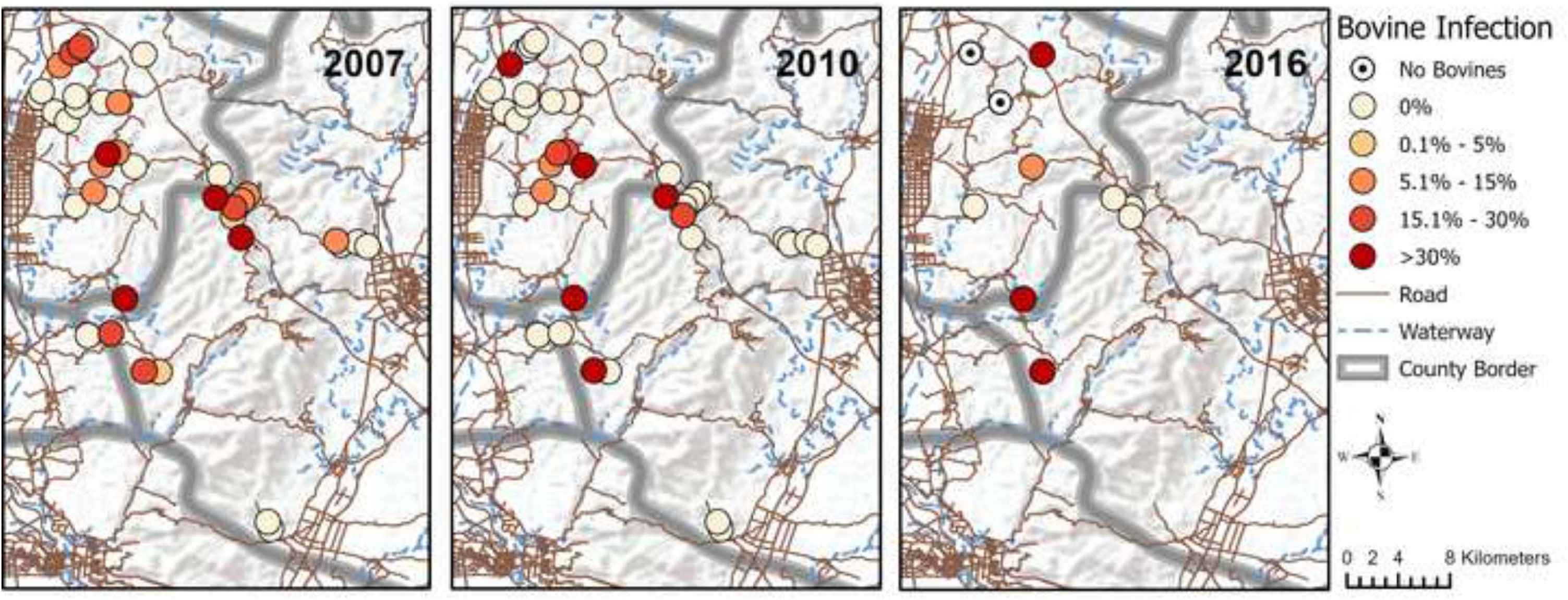
Village-level prevalence of schistosomiasis in bovines in 2007, 2010 and 2016. The prevalence of schistosomiasis infection in bovines is indicated for each village included in 2007 (left), 2010 (center) and 2016 (right). The darker the shade, the higher the prevalence. While villages in the lower left corner of each map were found to have repeatedly high infection prevalence, other villages achieved 0% infection in bovines in later years (upper left and center right of maps), while still others emerged as high prevalence villages in 2010 (center of maps) and 2016 (upper left of maps) after years of low infection prevalence in bovines. Service Layer Credits: World Terrain Base Sources: Esri, USGS, NOAA. OpenStreetMap Data Extracts for China, Asia: Data/Maps Copyright 2018 Geofabrik GmbH and OpenStreetMap Contributors. *Note: Figure 1 has a color version (online) and a grayscale version (print).*

The bovine study population was comprised of fewer water buffaloes than cattle (18.5% of all bovines were water buffalo in 2007; 15.5% in 2010), was predominantly female (86.3% in 2007; 87.2% in 2010), and ranged widely in age from less than a year to 26 years old.

Table 2 compares the distributions of each predictor variable by bovine infection status, for 2007, 2010 and 2016.

**Table 2.**
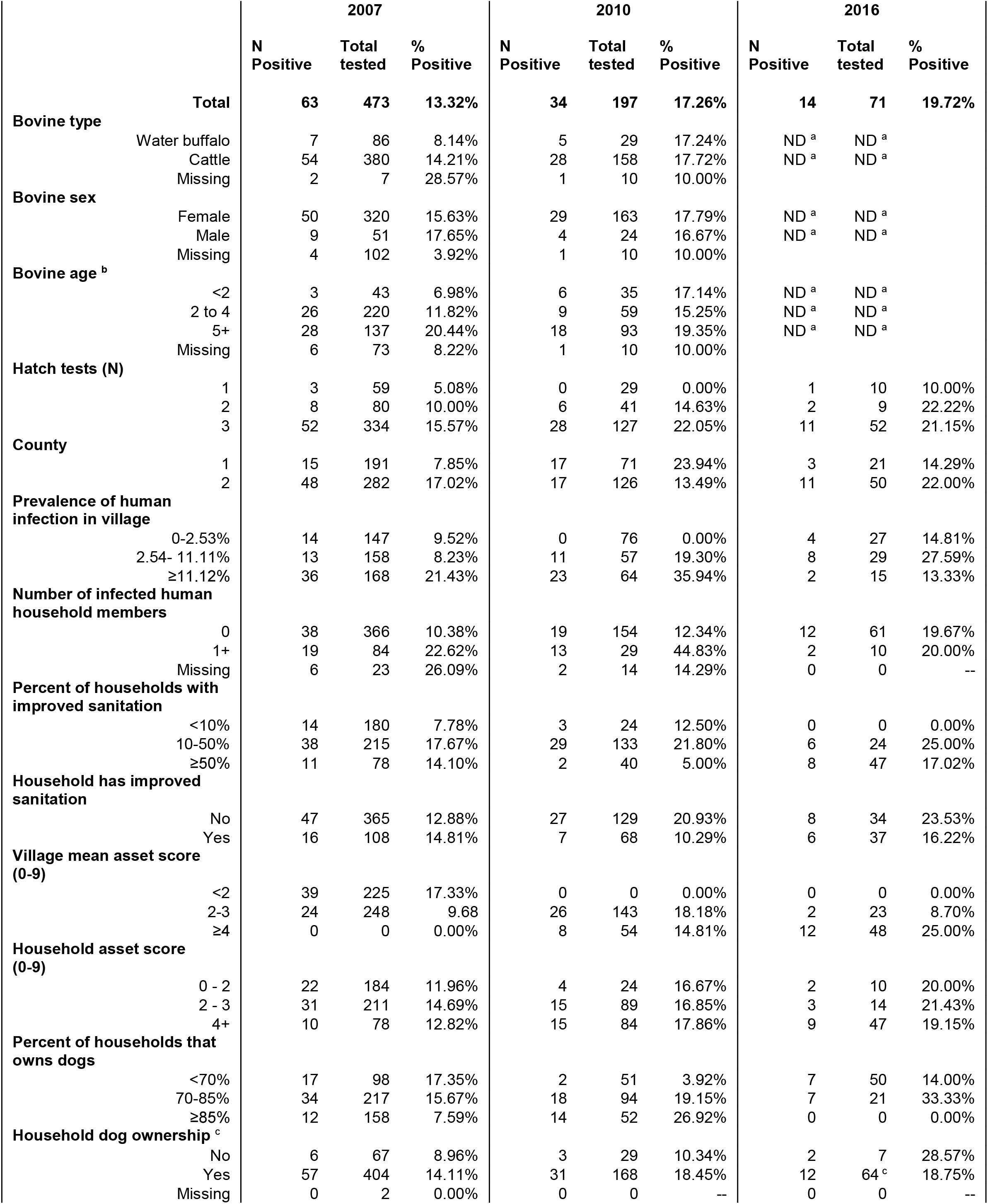
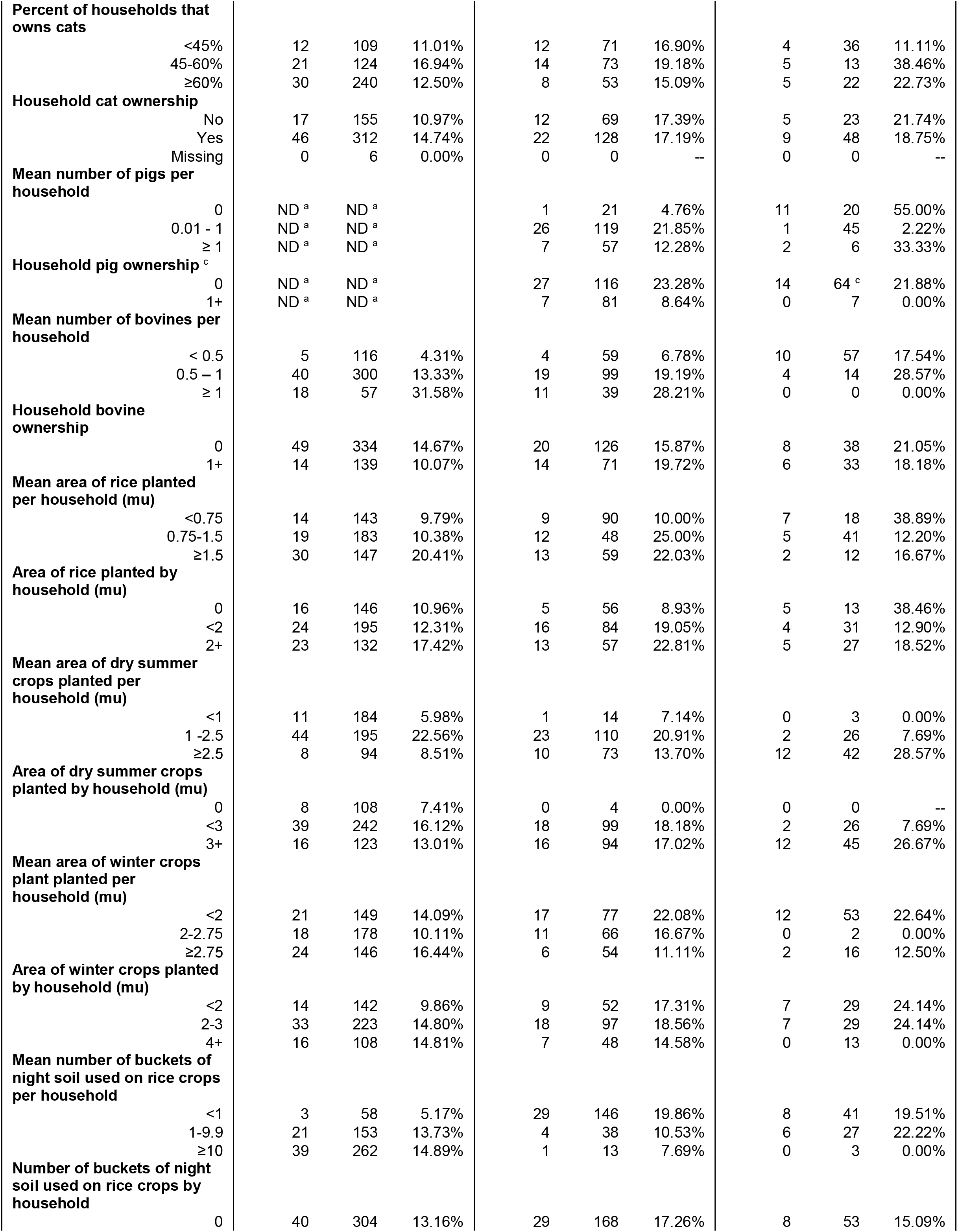
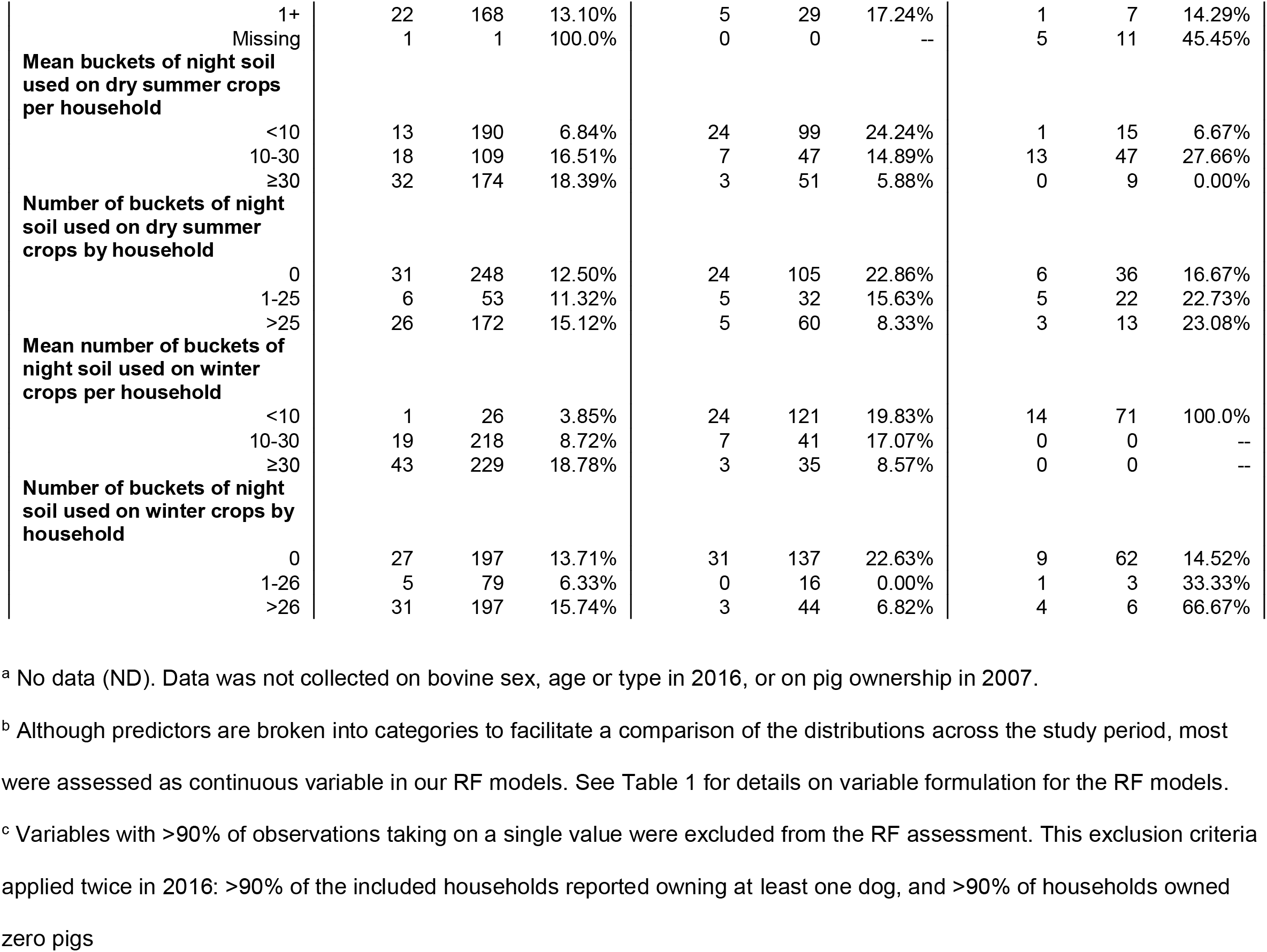
Tabulation of all predictors by bovine infection status.

There were several notable shifts in the distribution of predictors over time in our study region. Access to improved sanitation increased across our study period, rising from 22.8% of households reporting improved sanitation in 2007 to 52.1% with improved sanitation by 2016. Notably, the percent of bovines infected was roughly equal across the two sanitation groups in 2007, but by 2016, a greater proportion of bovines were infected if they belonged to households with unimproved sanitation (23.5%) compared to the households with improved sanitation (16.2%).Household assets also increased over time, as approximately half of all villages had a mean asset score of less than two in 2007, whereas in 2010 and 2016, none of the included villages had a mean asset score that low. Across the study period, there was a steady increase in the percent of households reporting planting rice (69.1% in 2007; 71.5% in 2010; 81.7% in 2016), and other summer crops (77.2% in 2007; 98% in 2010; 100% in 2016), as well as the cultivated area of rice and dry summer crops. Night soil use on rice and winter crops decreased over time: the proportion of households reporting any night soil use on rice crops dropped from 35.6% to 11.7% between 2007 and 2016, and for winter crops it dropped from 58.3% to 12.6%. By contrast, the proportion of night soil users for dry summer crops remained relatively constant across the study period (52.4% in 2007; 53.5% in 2010; 50.7% in 2016). Notably, the average amount of night soil being applied to crops dropped across the study period for all crop types.

### 3.2 Predictors of bovine infection

The predictor rankings were relatively stable across the full models, lean models and sensitivity models within each year, though there was more variability across collection years (Figure 2). For example, animal reservoirs were frequently among the top ten predictors in 2016, but only a household pig ownership (2010) and bovine density in the surrounding village (both 2007 and 2010) were among the top ten in earlier years.

**Figure 2.**
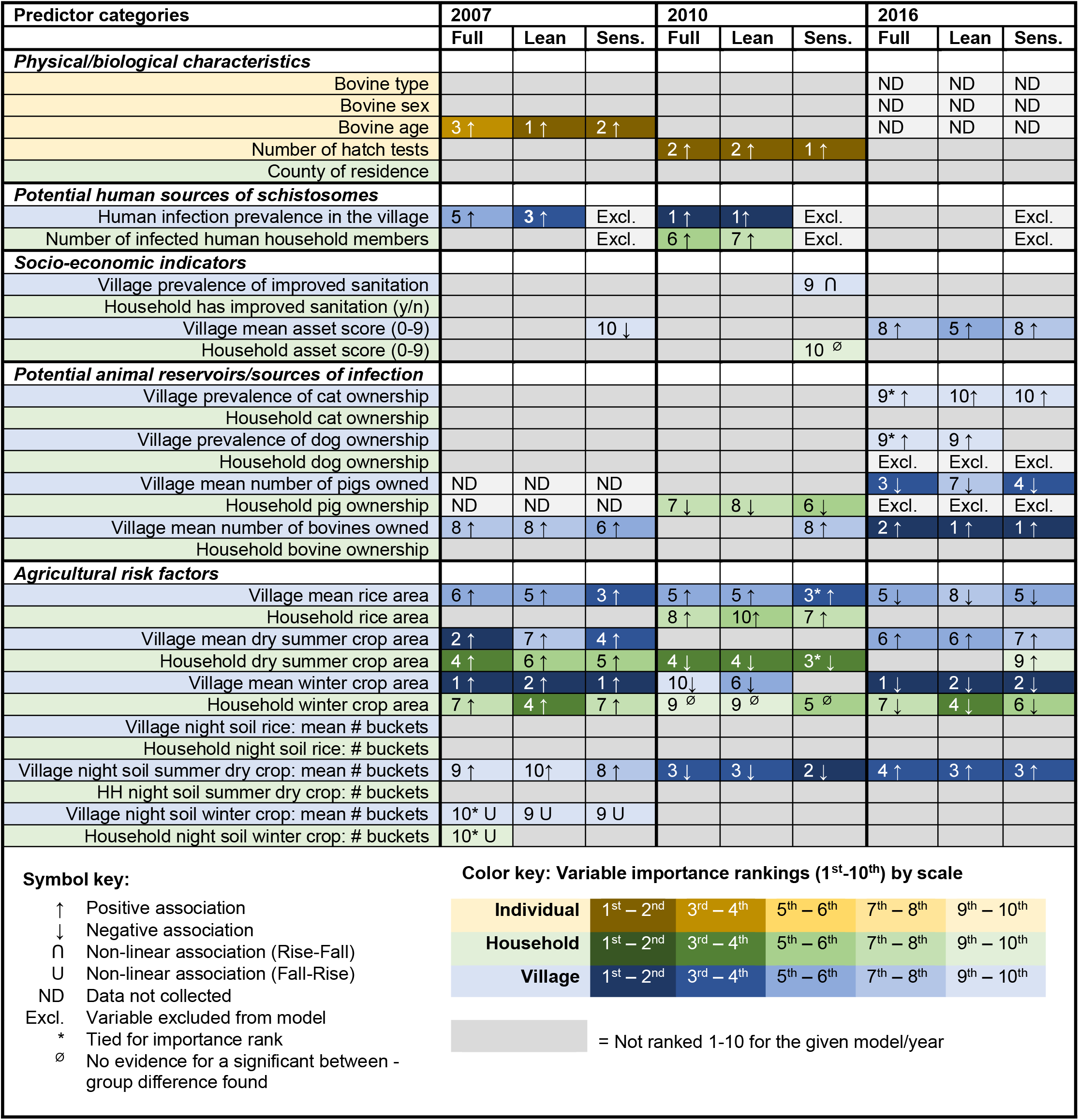
Variable importance rankings and direction of association for candidate predictors of bovine *S. japonicum* infection in 2007, 2010 and 2016. Variable importance rankings are based on a composite of mean decrease in accuracy scores for 10 random forest (RF) models for each model type (full, lean and sensitivity) and collection year. The direction of association was determined through logistic regression, using tertile categories for continuous variables to assess evidence for non-linearity. A p-value of <0.2 was used to indicate evidence of a between-group difference, and, when a between group difference was found, the direction of association is indicated. See Supplementary Table S2 for detailed logistic regression results. *Note: Figure 2 will be in color for both the online and print versions.*

The density of bovine in a village was one of the most consistent predictors: it was scored in the top ten in at least one RF analysis from each year, all of which indicate that an increase in bovine density in the surrounding village corresponds with an increase in bovine infection risk. Pig ownership was associated with a decrease in bovine infection risk in both years that it was assessed.

Agricultural practices was the most frequently ranked predictor category across all years. Specifically, the household area of winter crops planted, the mean area of rice planted in the surrounding village, and the mean amount of night soil applied to dry summer crops in the surrounding village were all among the top ten predictors in all models. Additionally, the total household area of summer crops planted and the village mean area of winter crops were also all among the top ten predictors in at least one of the three model types for 2007, 2010 and 2016. However, despite the inter-year agreement indicating the importance of agricultural predictors, the direction of association between the top agricultural predictors and bovine infection was not always consistent across years. In 2007, increases in all key agricultural predictors were associated with an increase in bovine infection risk, apart from night soil use on winter crops. By contrast, in 2010 and 2016 our models indicate a mixture of positive and negative associations across the key agricultural predictors, and in one instance (household winter crop area in 2010), no evidence of a relationship was found.

Figure 3 depicts changes in the distribution of different agricultural practices by bovine infection status between 2007 and 2016. Most prominent among these is dry summer crop farming, which saw a notable increase in the total and mean area of crop being planted by households and villages between 2007 and 2016 (Figure 3, panel B). In 2007 and 2016 the area of dry summer crops cultivated at the household and village-level was positively associated with bovine infection, but in 2010 it was not. There was a general decrease in the amount of night soil being applied to rice, winter crops and, to a lesser extent, dry summer crops, between 2007 and 2016 (Figure 3, panels D-F). Greater night soil use on dry summer crops at the village level was associated with increased bovine infection risk in 2007 and 2016, but not in 2010. Less night soil was used on rice, compared to other crops, and night soil use on rice was not a key predictor of bovine infection in any year.

**Figure 3.**
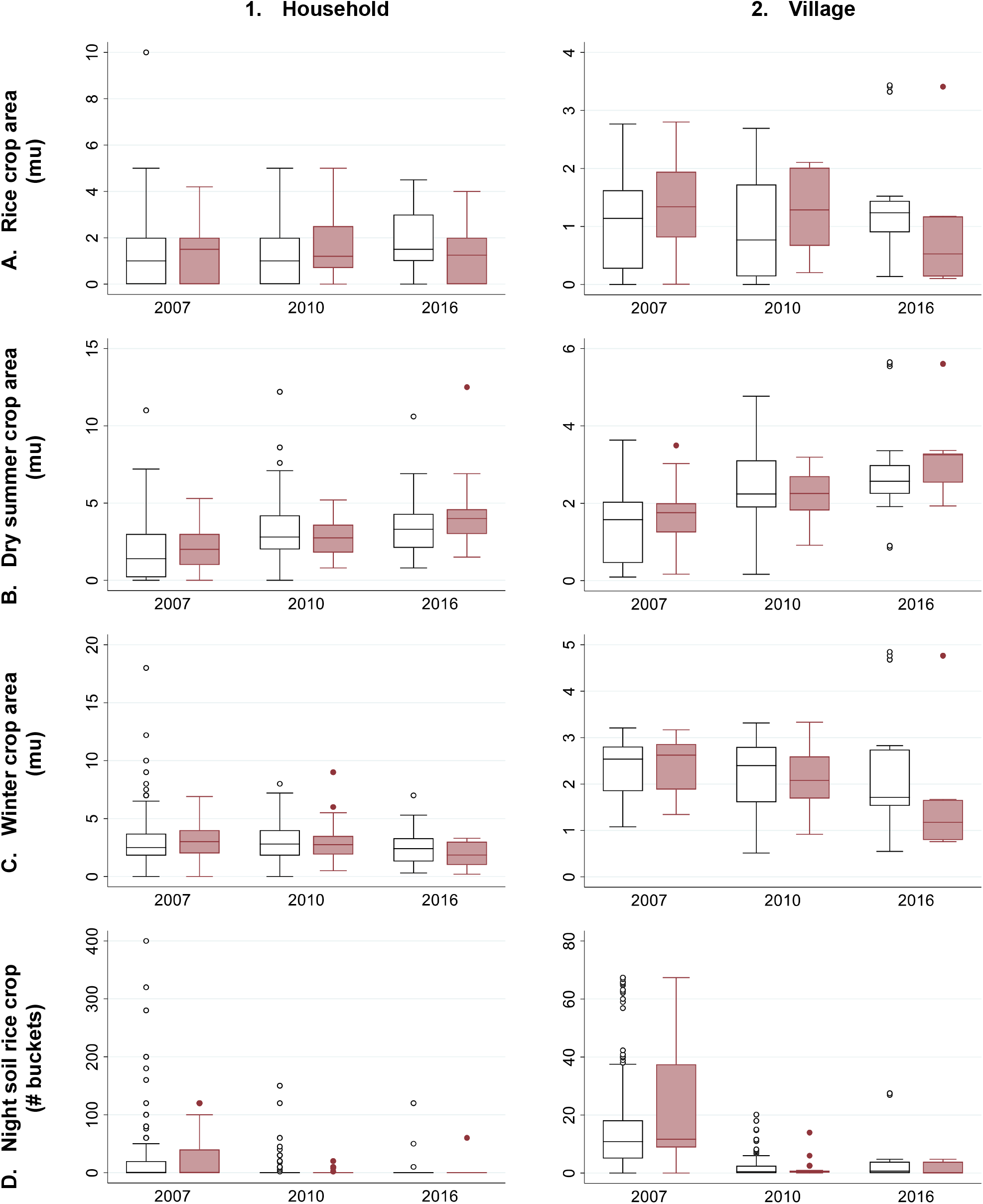
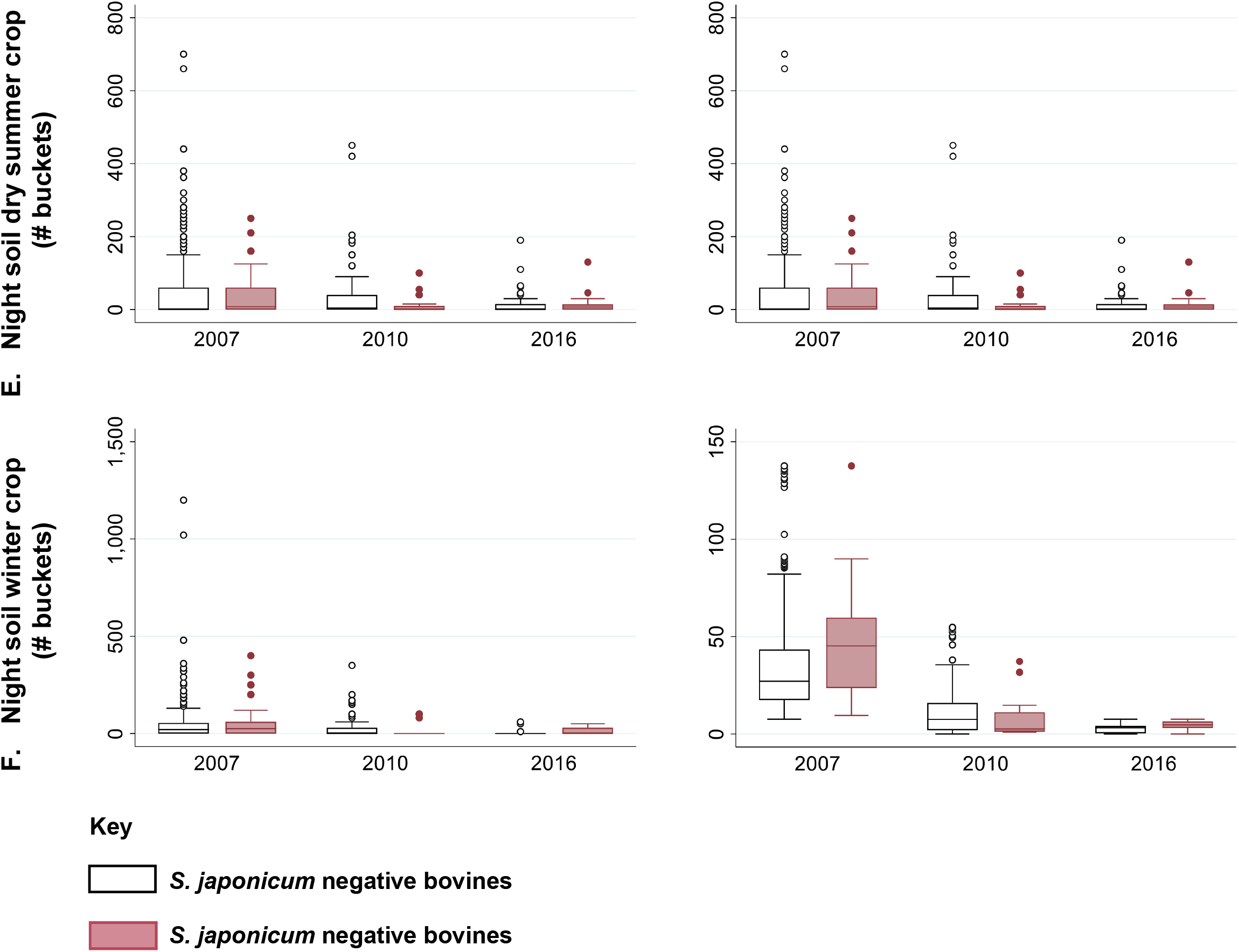
Changes in agricultural practices and the relationship between bovine infection and agricultural predictors over time. For each of the agricultural predictors included in this analysis, boxplots are used to represent the distribution of uninfected (white) and infected bovines, for household-level (left) and village-level variables (right) in 2007, 2010 and 2016. *Note: Figure 3 has a color version (online) and a grayscale version (print).*

There are several other key predictors that stand out in one or more collection years. Human infection prevalence in the surrounding village was among the top five predictors of bovine infection in 2007 and 2010, and the number of infected humans within the household was among the top ten predictors in 2010. In all cases, an increase in human infections was associated with an increase in bovine infections. When the human infection predictors were removed for the sensitivity analysis, the rankings of the remaining predictors did not shift substantially in any collection year. Bovine age was also among the top predictors in 2007, and the probability of infection was found to increase with age. In all of the 2010 analyses, the number of hatch tests was an important predictor of bovine infection, a feature not shared by the 2007 and 2016 analyses. This may be related to the lower proportion of bovines with three hatch test results in 2010 (64.5%), as compared to 2007 (70.6%) and 2016 (73.2%).

Of the three different analyses performed (full, lean and sensitivity) for each collection year, the full models (i.e. those that included the full list of predictors available in a given year) tended to perform the best (Table 3, Figure 4). Overall, the models had high accuracy values, with the top performing full models producing a maximum accuracy of 0.864 (95% CI: 0.79 − 0.92) in 2007, 0.816 (95% CI: 0.68 − 0.91) in 2010, and 1.0 (0.81 − 1.0) in 2016. Furthermore, despite some variation in model performance across the ten iterations of RF models for each analysis year, ultimately the models were relatively stable. For example, across each of the ten iterations of full model analyses, the AUC range was relatively narrow: 0.724 − 0.75 in 2007, 0.816 − 0.863 in 2010, and 0.982 1.0 in 2016. However, due to class imbalance in the reserved test datasets (see the no information rate (NIR) in Table 3), the Kappa value is also a useful performance metric for our models, as this takes class imbalance into account. According to the benchmarks laid out by Landis and Koch (1977), the Kappa statistics from our 2007 analyses suggest a “Fair” level of agreement (0.21 − 0.40) between our best RF models and the true known values in 2007. For 2010, the highest Kappa statistic came from the full predictor analysis, with a Kappa of 0.463, indicating a “Moderate” level of agreement (0.41 − 0.60) between the prediction model and the reserved test dataset (Landis and Koch, 1977). In 2016, both the full and sensitivity models achieved perfect prediction (Kappa = 1) for the test dataset in at least one of the ten model iterations, whereas the Kappa statistic for the top performing lean model was 0.853, or “Almost Perfect”, according to Landis & Koch (Landis and Koch, 1977).

**Table 3.**
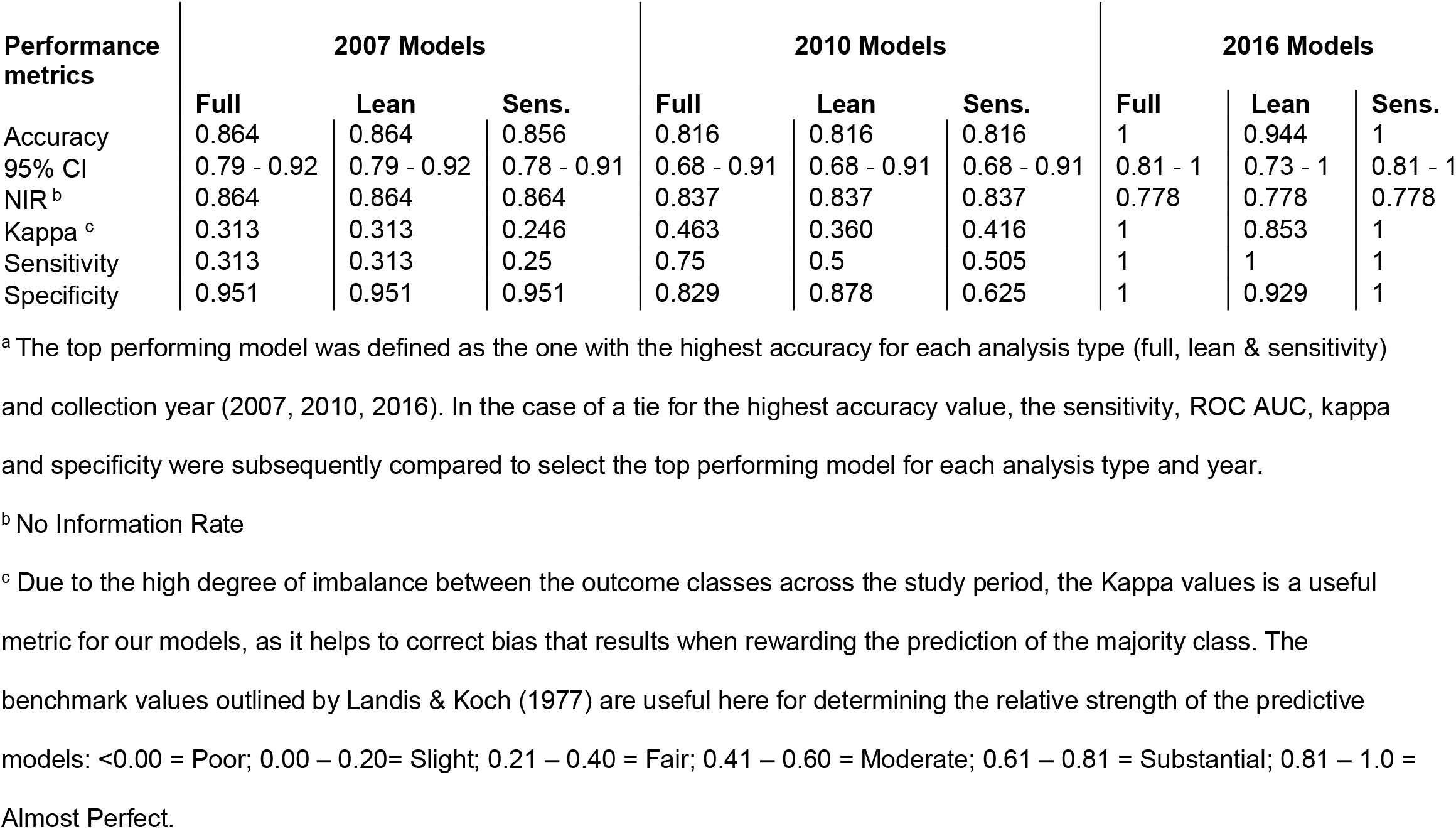
Comparison of model performance metrics for the top performing model ^a^ from the full, lean and sensitivity analyses in 2007, 2010 and 2016.

| Performance metrics | 2007 Models |  |  | 2010 Models |  |  | 2016 Models |  |  |
| --- | --- | --- | --- | --- | --- | --- | --- | --- | --- |
|  | Full | Lean | Sens. | Full | Lean | Sens. | Full | Lean | Sens. |
| Accuracy | 0.864 | 0.864 | 0.856 | 0.816 | 0.816 | 0.816 | 1 | 0.944 | 1 |
| 95% CI | 0.79 - 0.92 | 0.79 - 0.92 | 0.78 - 0.91 | 0.68 - 0.91 | 0.68 - 0.91 | 0.68 - 0.91 | 0.81 - 1 | 0.73 - 1 | 0.81 - 1 |
| NIR <sup>b</sup> | 0.864 | 0.864 | 0.864 | 0.837 | 0.837 | 0.837 | 0.778 | 0.778 | 0.778 |
| Kappa <sup>c</sup> | 0.313 | 0.313 | 0.246 | 0.463 | 0.360 | 0.416 | 1 | 0.853 | 1 |
| Sensitivity | 0.313 | 0.313 | 0.25 | 0.75 | 0.5 | 0.505 | 1 | 1 | 1 |
| Specificity | 0.951 | 0.951 | 0.951 | 0.829 | 0.878 | 0.625 | 1 | 0.929 | 1 |

<sup>a</sup> The top performing model was defined as the one with the highest accuracy for each analysis type (full, lean & sensitivity) and collection year (2007, 2010, 2016). In the case of a tie for the highest accuracy value, the sensitivity, ROC AUC, kappa and specificity were subsequently compared to select the top performing model for each analysis type and year.
<sup>b</sup> No Information Rate
<sup>c</sup> Due to the high degree of imbalance between the outcome classes across the study period, the Kappa values is a useful metric for our models, as it helps to correct bias that results when rewarding the prediction of the majority class. The benchmark values outlined by Landis & Koch (1977) are useful here for determining the relative strength of the predictive models: <0.00 = Poor; 0.00 – 0.20= Slight; 0.21 – 0.40 = Fair; 0.41 – 0.60 = Moderate; 0.61 – 0.81 = Substantial; 0.81 – 1.0 = Almost Perfect.

**Figure 4.**
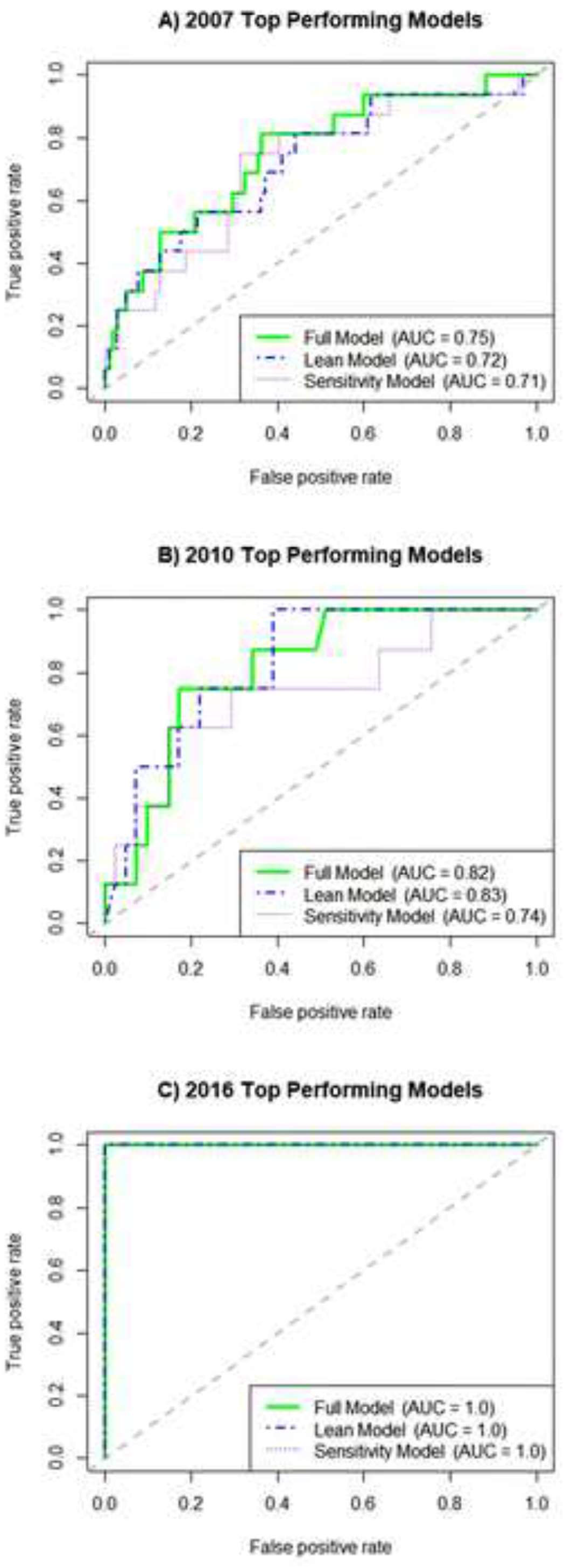
The ROC curves and corresponding AUC are shown for the top performing full, lean and sensitivity models for A) 2007 in the top panel, B) 2010 in the middle panel, and C) 2016 in the bottom panel. *Note: Figure 4 has a color version (online) and a grayscale version (print).*

Because the full list of predictors changed slightly across the collection years, a supplemental analysis was conducted in which only predictors that were available in all three collection years were included in the RF models. This analysis demonstrated that 1) intra-year rankings and extra-year patterns did not change substantially, and 2) agricultural variables remained the most prominent predictor category when comparing across the entire study period. See Supplementary Figure S1 for details of the supplemental analysis. **4**.

## Discussion

Agricultural risk factors, bovine density, human infection, pig ownership and bovine age were all found to be key predictors of bovine infection status in one or more of the years assessed in this analysis. Of the agricultural risk factors, night soil use on dry summer crops, the village-level area of rice crops, and both the household and village-level areas of dry summer and winter crops were each ranked among the top five predictors for one or more collection years in our RF models. Interestingly, for 2007, all of the ranked agricultural variables except night soil use on winter crops were associated with an increase in bovine infection risk in our logistic regression assessments, whereas in 2010 and 2016, these agricultural factors were found to be variably positively and negatively associated with infection. For example, the area of rice cultivated in a village was positively associated with bovine infection in 2007 and 2010, but negatively associated in 2016. This finding may be related to changing norms and interventions that have taken hold in recent years as a result of increasing awareness of the potential risks posed by both bovines as a reservoir of schistosomiasis, and specific agricultural practices. For example, across our study period, we saw a steady increase in the percent of households planting rice (69.1% in 2007; 71.5% in 2010; 81.7% in 2016), and a simultaneous decrease in the percent of households applying any night soil to their rice crops (35.6% in 2007, 14.7% in 2010; 11.7% in 2016). These shifting norms in rice production and night soil use likely resulted in a decrease in the overall concentration of night soil on rice crops within our study villages, which in turn, may help to explain why the village-level rice crop area shifts from having a positive association with bovine infection in 2007 and 2010, to a negative association by 2016.

Assessments conducted in China early in the new millennium repeatedly highlighted bovines as a key source of environmental contamination and as the main animal reservoir of *S. japonicum* in the country (Gray et al., 2009a; Gray et al., 2007; Guo et al., 2006). Beginning in 2004, a new government-led approach to eliminating schistosomiasis transmission in China was adopted, which − in conjunction with infrastructure improvements in rural areas and several new schistosomiasis elimination interventions − featured replacing bovines with machinery in agricultural production (Liu et al., 2017). Thus, the negative associations that were found intermittently between bovine infection and some of our agricultural variables in 2010 and 2016 may be linked to the added precautions that were being adopted when bovines were being used for agriculture, or because bovines were being reallocated for other purposes (e.g. beef production) as machinery became the norm for large crop areas or those deemed high risk (e.g. wet rice crops).

Dry summer crops stand out as a key agricultural risk factor for bovine infection. The area of dry summer crops cultivated and the quantity of night soil applied to dry summer crops were key predictors of bovine infection in all years, and positively associated with bovine infection in 2007 and 2016. It is unclear why this is: it is possible that the cultivation of such crops provides snail habitat and/or the use of night soil on dry summer crops facilitates infection of snails living near the field (in our study region, more night soil is applied to dry summer crops than rice), leading to bovine infection. A downward trend in night soil use (total and mean number of buckets) on crops can be observed in Figure 3 (parts D-F), though notably, we do not see any substantial shift in the overall proportion of households that reported any night soil use on summer crops over the years (52.4% in 2007; 53.5% in 2010; 50.7% in 2016) (Table 2). Increasing recognition of the potential risks posed by night soil use during our study period (Carlton et al., 2011) may have also contributed to some decreases in night soil use and/or the more careful treatment of night soil prior to field applications. However, the continued practice of applying night soil to dry summer crops, paired with the steady increase in the total area of summer crops being planted by villagers over the study period (see Figure 3, part B) may help to explain why night soil use on summer crops is positively association with bovine infection status in 2016.

Bovine density in the surrounding village was in the top ten predictors of our RF models, and was positively associated with bovine infection in our regression models in all collection years. These findings align well with the existing literature that points to bovines as the most important reservoir of *S. japonicum* infection in China (Gray et al., 2009a), and suggests that being in close proximity to higher densities of bovine hosts may correspond with increasing infection risk, as has been found for other bovine pathogens (Guo et al., 2006; Meadows et al., 2018; Spencer et al., 2015). However, it is worth noting that household-level bovine ownership was not among the top predictors in any of our RF models, highlighting that the larger-scale lens (i.e. village-level analysis scale) may be particularly important to future investigations and control strategies. Likewise, recent informal interviews with residents in our study sites have revealed that bovines are infrequently kept near the home, as allowing bovines to graze (and defecate) freely is an economical and efficient way of raising bovines, further illustrating that the household scale may not always be broad enough to capture larger scale trends. Instead, villagers opt to bring their bovines to the mountains to graze during the day, which subsequently presents more opportunities for contact between bovines from different households, and may ultimately result in more widespread environmental contamination (e.g. bovine feces washed into nearby irrigation ditches after precipitation).

In the developmental stages of this analysis, we hypothesized that human infection prevalence and the number of infected people in the household would be among the top predictors of bovine infection status, given the known link between human schistosomiasis and bovine reservoirs (e.g. Gray et al., 2009a). It was therefore somewhat surprising to find that household-level human infection was only highly ranked in 2007 within our RF models, while village human infection prevalence was ranked in the top ten in 2007 and 2010, and neither household-level human infection nor village-level human infection prevalence were important predictors in 2016. One potential explanation for the changing importance of human infection as a predictor of bovine infection could be related to the aforementioned bovine-removal phenomenon, in which bovines are increasingly being removed from the village area and brought to alternative mountain locations for grazing. As this act would likely result in an<u>in an</u> overall drop in time humans spent in bovine grazing areas, one might expect to see a separation of the bovine and human infection cycles if snail habitats are common in bovine grazing sites. In fact, the drop in the important rankings of human infection status in 2016 coincides with a jump in the variable importance rankings for village-level bovine ownership (6^th^ − 8^th^ in 2007 and 2010; 1^st^ -2^nd^ in 2016), providing further support of the theory that bovines may be becoming increasing important reservoirs of schistosomiasis infection. On the other hand, an altogether different explanation for the differences in the 2016 rankings compared to 2007 and 2010 is that the 2016 data collection simply didn’t have a large enough sample size to allow for the detection of a true relationship between relatively rare events.

As such, one limitation of this assessment was the relatively small sample sizes, particularly in 2016 (N=71), though to a lesser extent, 2010 (N=197) and 2007 (N=473), given the correspondingly large number of predictors that were included in the full predictor models (N=29, N=31, N=26, in 2007, 2010 and 2016 respectively). While RF models are generally acknowledged as being able to handle assessments of high dimensional data even with relatively small sample sizes (Biau and Scornet, 2016), it remains that small samples sizes can still give rise to the aforementioned issue of non-detection of rare events. Another limitation to this assessment is that RF models tends to favor continuous predictors over categorical measures, as they allow for a wider range of potential split points for classifying observations. For this reason, it is not particularly surprising that age was the only predictor from the individual/physical characteristics predictor group that was ranked among the top ten predictors, as the remaining individual characteristics were binary measures. Another notable limitation of the variable importance rankings used in RF models is that they become less reliable when predictors are highly correlated with one another (Strobl et al., 2008). This may be particularly important to the rankings ascribed to the agricultural variables, as correlation between the area of the different crop types planted and the amount of night soil used on each crop tended to be high across all collection years, with the highest predictor correlations found in the 2016 collection year (See Supplementary Figures S2 − S4 for correlation matrices). This is notable, as a higher degree of instability in the variable importance rankings was also found for 2016 as compared to 2010 or 2007, suggesting predictor correlation may be responsible. We therefore recommend that the variable rankings presented in this analysis be interpreted more holistically (e.g. agricultural variables are strong predictors of bovine infection), and advise caution when comparing unique variable ranking values against one another (e.g. rice crop area is less important than winter crop area).

Our main interests in this assessment were to 1) identify the best physical and environmental predictors of bovine *S. japonicum* infection within rural farming communities in Sichuan China, and 2) to ascertain whether there are broader trends in bovine infection distribution across individual, household or village-levels scales or over time. Our RF assessments have highlighted some key patterns that were repeated across multiple collection years and multiple iterations of three different models. Agricultural factors and animal reservoirs − specifically, high bovine density in the surrounding village − were repeatedly found to be among the top predictors of bovine *S. japonicum* infection across all assessment years. Furthermore, human infection, pig ownership and bovine age were also found to be strong predictors of bovine infection in at least one year. Taken together, these findings highlight the potential utility of presumptively treating bovines residing in villages and households that engage in high-risk agricultural practices, those of older age, or those belonging to villages with particularly high human infection or bovine density. Additionally, village-level predictors tended to be better predictors of bovine infection than household-level predictors, suggesting that future investigations and interventions may need to apply a broad ecological lens in order to successfully extricate and address environmental sources of ongoing transmission. In so doing, not only can unnecessary bovine morbidity and the associated economic losses be reduced, but such measures will also help to minimize the potential for bovines to serve as another pathway to persistent human schistosomiasis transmission.

## Data Availability

Due to the inclusion of potentially identifying information (e.g. socio-economic indicators and human infection status), access to study data must be requested through the Carlton Lab group to ensure the protection of research subjects and compliance with all ethical guidelines. Please contact Elizabeth Carlton at for more information.

## Acknowledgments

We are grateful for the support and efforts of the field research team members from the Institute of Parasitic Diseases and the county anti-schistosomiasis control stations for their efforts in collecting the data presented here.

## Author Contributions

Conceptualization: EC, YL, SP, AB; Data Curation: DL, EG; Formal Analysis: EG, SP, KK, AB, EC; Funding Acquisition: EC, YL, SP; Investigation: YL, DL, EC; Methodology: EG, SP, EC, AB, KK; Project Administration: YL, DL; Resources: EC, YL, DL; Validation: EG, SP, EC, KK; Writing − Original Draft Preparation: EG, SP, EC, KJ; Writing − Review & Editing: EG, SP, KK, YL, DL, EC, AB, KJ.

