## Supplementary Materials for "Predictors of bovine *Schistosoma japonicum* infection in rural Sichuan, China"

**Supplementary Data**

**Supplementary Table S1. Village, household and bovine inclusion.** Village inclusion required that at least one household owned bovines, and that at least one bovine was tested for schistosomiasis infection within the village. Participating households were defined as those with both household survey and infection survey data for one or more bovine. Thus, bovines were ultimately included in this analysis if both infection surveys and household surveys were performed for the given bovine.

|  | **2007** | **2010** | **2016** |
| --- | --- | --- | --- |
|  | N | N | N |
| **Village inclusion** |  |  |  |
| Number of villages surveyed | 36 | 36 | 10 |
| Number of villages with households that owned bovines | 36 | 35 | 8 |
| Total number of villages with bovine infection data | 35 | 30 | 8 |
| Total number of villages included ^a^ | 35 | 30 | 8 |
| **Household inclusion** |  |  |  |
| Total number of households in included villages | 1525 | 823 | 400 |
| Number of households reporting owning bovines from included villages | 529 | 247 | 65 |
| Number of households with bovine infection data from included villages | 432 | 211 | 58 |
| Total number of households included in this analysis ^a^ | 403 | 178 | 57 |
| **Bovine inclusion** |  |  |  |
| Total number of bovines reported on household surveys in included villages | 711 | 335 | 76 |
| Number of bovines with infection data from included villages | 503 | 233 | 72 |
| Total number of bovines included in this analysis ^a^ | 473 | 197 | 71 |

^a^ The total number of included villages, households and bovines was determined by excluding those that didn’t have any bovines with household survey data (i.e. the predictor variables) and those that didn’t have any bovine infection survey data (i.e. the outcome variable).

**S2 Table. Simple logistic regression analyses to determine the direction of association between bovine infection status and each predictor, by collection year.** Tertiles (and sometimes quartiles) by year were used in simple logistic regression analyses to help investigate potential non-linearity. All predictor that were given a final rank of 1^st^ – 10^th^ in one of the RF models (full, lean, sensitivity) for a given year were examined for between-group differences.

|  | **2007** | | | | **2010** | | | | **2016** | | | |
| --- | --- | --- | --- | --- | --- | --- | --- | --- | --- | --- | --- | --- |
|  | **Tertile** | **PE ^a^** | **SE** | **P-value** | **Tertile** | **PE ^a^** | **SE** | **P-value** | **Tertile** | **PE ^a^** | **SE** | **P-value** |
| ***Individual characteristics*** |  |  |  |  |  |  |  |  |  |  |  |  |
| Bovine age |  |  |  |  |  |  |  |  |  |  |  |  |
|  | ≤ 3 | ref |  |  |  |  |  |  |  |  |  |  |
|  | 3.1 - 5 | 0.58 | 0.34 | 0.094* |  |  |  |  |  |  |  |  |
|  | ≥ 5.1 | 0.92 | 0.36 | 0.011* |  |  |  |  |  |  |  |  |
| # of hatch tests |  |  |  |  |  |  |  |  |  |  |  |  |
|  |  |  |  |  | 1 | p.f.p | p.f.p | p.f.p.^b,^ * |  |  |  |  |
|  |  |  |  |  | 2 | Ref |  |  |  |  |  |  |
|  |  |  |  |  | 3 | 0.50 | 0.49 | 0.308 |  |  |  |  |
| ***Human sources*** |  |  |  |  |  |  |  |  |  |  |  |  |
| Human infection: household count |  |  |  |  |  |  |  |  |  |  |  |  |
|  |  |  |  |  | 0 | Ref |  |  |  |  |  |  |
|  |  |  |  |  | 1+ | 1.75 | 0.45 | <0.001* |  |  |  |  |
| Human infection: village prevalence |  |  |  |  |  |  |  |  |  |  |  |  |
|  | ≤ 2.6 | Ref |  |  | 0 | p.f.p. | p.f.p | p.f.p.^b,^ * |  |  |  |  |
|  | 2.7 - 11.9 | -0.24 | 0.40 | 0.558 | 0.1 – 10.8 | Ref |  |  |  |  |  |  |
|  | ≥ 12 | 1.06 | 0.33 | 0.001* | ≥ 10.9 | 1.11 | 0.43 | 0.010* |  |  |  |  |
| ***Socio-economic*** |  |  |  |  |  |  |  |  |  |  |  |  |
| Vil: Imp. sanitation |  |  |  |  | ≤ 25 | Ref |  |  |  |  |  |  |
|  |  |  |  |  | 26 – 42 | 0.87 | 0.43 | 0.046* |  |  |  |  |
|  |  |  |  |  | ≥ 43 | -0.99 | 0.62 | 0.112* |  |  |  |  |
| Vil. asset score (0-9) |  |  |  |  |  |  |  |  |  |  |  |  |
|  | ≤ 1.65 | Ref |  |  |  |  |  |  | ≤ 4.03 | Ref |  |  |
|  | 1.66 – 2.5 | -1.09 | 0.35 | 0.002* |  |  |  |  | 4.04 - 4.59 | 1.40 | 0.86 | 0.104* |
|  | ≥ 2.51 | -1.15 | 0.34 | <0.001* |  |  |  |  | ≥ 4.6 | 1.23 | 0.90 | 0.170* |
| HH asset score (0-9) |  |  |  |  |  |  |  |  |  |  |  |  |
|  |  |  |  |  | ≤ 2 | Ref |  |  |  |  |  |  |
|  |  |  |  |  | 3 | 0.32 | 0.51 | 0.531 |  |  |  |  |
|  |  |  |  |  | 4 | 0.04 | 0.59 | 0.951 |  |  |  |  |
|  |  |  |  |  | ≥ 5 | 0.41 | 0.53 | 0.440 |  |  |  |  |
| ***Animal reservoir*** |  |  |  |  |  |  |  |  |  |  |  |  |
| Vil.: cat ownership |  |  |  |  |  |  |  |  | ≤ 43.9 | Ref |  |  |
|  |  |  |  |  |  |  |  |  | 44 – 58.9 | 2.81 | 1.11 | 0.012* |
|  |  |  |  |  |  |  |  |  | ≥ 59 | 2.07 | 1.14 | 0.069* |
| Vil.: dog ownership |  |  |  |  |  |  |  |  | ≤ 64 | Ref |  |  |
|  |  |  |  |  |  |  |  |  | 64.1 – 74.9 | 0.24 | 0.82 | 0.769 |
|  |  |  |  |  |  |  |  |  | ≥ 75 | 1.25 | 0.77 | 0.104* |
| Vil. mean pigs owned |  |  |  |  |  |  |  |  | ≤ 0.14 | Ref. |  |  |
|  |  |  |  |  |  |  |  |  | 0.21 – 0.43 | -2.05 | 1.09 | 0.060* |
|  |  |  |  |  |  |  |  |  | ≥ 0.44 | -1.30 | 0.83 | 0.119* |
| HH: pigs owned |  |  |  |  | 0 | Ref |  |  |  |  |  |  |
|  |  |  |  |  | 1 | -0.42 | 0.67 | 0.534 |  |  |  |  |
|  |  |  |  |  | >1 | -1.50 | 0.56 | 0.008* |  |  |  |  |
| Vil.: mean bovines owned | ≤ 0.65 | Ref |  |  | ≤ 0.55 | Ref |  |  | ≤ 0.30 | Ref |  |  |
|  | 0.66 – 0.89 | 1.19 | 0.38 | 0.002* | 0.56 – 0.74 | -0.10 | 0.51 | 0.839 | 0.30 – 0.44 | 0.63 | 0.96 | 0.513 |
|  | ≥ 0.89 | 1.16 | 0.40 | 0.003* | ≥ 0.75 | 0.77 | 0.45 | 0.086* | ≥ 0.441 | 2.24 | 0.86 | 0.009* |
| ***Agriculture*** |  |  |  |  |  |  |  |  |  |  |  |  |
| Vil. mean rice area |  |  |  |  |  |  |  |  |  |  |  |  |
|  | ≤ 0.84 | Ref |  |  | ≤ 0.61 | Ref |  |  | ≤ 0.95 | Ref |  |  |
|  | 0.84 – 1.47 | -0.33 | 0.37 | 0.380 | 0.62 – 1.29 | 2.33 | 0.77 | 0.003* | 0.96 – 1.34 | -0.97 | 0.70 | 0.166* |
|  | ≥ 1.47 | 0.55 | 0.32 | 0.081* | ≥ 1.30 | 2.35 | 0.77 | 0.002* | ≥ 1.35 | -1.61 | 0.86 | 0.061* |
| HH rice area |  |  |  |  |  |  |  |  |  |  |  |  |
|  |  |  |  |  | ≤ 0.5 | Ref |  |  |  |  |  |  |
|  |  |  |  |  | 0.6 – 1.5 | 0.57 | 0.49 | 0.253 |  |  |  |  |
|  |  |  |  |  | ≥ 1.6 | 0.76 | 0.48 | 0.116* |  |  |  |  |
| Vil sum. crop area |  |  |  |  |  |  |  |  |  |  |  |  |
|  | ≤ 0.73 | Ref |  |  |  |  |  |  | ≤ 2.412 | Ref |  |  |
|  | 0.74 – 1.96 | 1.51 | 0.39 | <0.001* |  |  |  |  | 2.413 – 2.98 | 0.88 | 0.92 | 0.336 |
|  | ≥ 1.97 | 0.90 | 0.42 | 0.031* |  |  |  |  | ≥ 2.99 | 1.81 | 0.86 | 0.034* |
| HH sum. crop area |  |  |  |  |  |  |  |  |  |  |  |  |
|  | ≤ 1 | Ref |  |  | ≤ 2.2 | Ref |  |  | ≤ 2.8 | Ref |  |  |
|  | 1.1 – 2.2 | 0.69 | 0.34 | 0.041* | 2.3 – 3.8 | -0.09 | 0.43 | 0.830 | 2.81 – 4.1 | 1.70 | 0.86 | 0.046* |
|  | ≥ 2.3 | 0.40 | 0.33 | 0.230 | ≥ 3.9 | -0.69 | 0.50 | 0.167* | ≥ 4.2 | 0.84 | 0.92 | 0.362 |
| Vil. win. crop area* |  |  |  |  |  |  |  |  |  |  |  |  |
|  | ≤ 1.869 | Ref |  |  |  |  |  |  |  |  |  |  |
|  | 1.87 – 2.55 | 0.01 | 0.40 | 0.981 | ≤ 1.869 | Ref |  |  | ≤ 1.58 | Ref |  |  |
|  | 2.56 – 2.81 | 0.01 | 0.40 | 0.981 | 1.87 – 2.58 | -0.50 | 0.45 | 0.269 | 1.59 – 1.89 | -0.86 | 0.70 | 0.219 |
|  | ≥2.82 | 0.48 | 0.37 | 0.193* | ≥ 2.59 | -0.60 | 0.46 | 0.193* | ≥ 1.9 | -1.55 | 0.86 | 0.071* |
| HH win. crop area* |  |  |  |  |  |  |  |  |  |  |  |  |
|  | 1.79 | Ref |  |  | ≤ 1.9 | Ref |  |  | ≤ 1.2 | Ref |  |  |
|  | 1.8 –2.5 | -0.03 | 0.41 | 0.944 | 2 – 2.8 | -0.07 | 0.53 | 0.895 | 1.3 – 2.1 | -0.62 | 0.81 | 0.442 |
|  | 2.6 – 3.7 | 0.54 | 0.38 | 0.152* | 2.9 – 3.7 | 0.36 | 0.51 | 0.617 | 2.2 – 3.2 | -0.76 | 0.80 | 0.340 |
|  | ≥ 3.8 | 0.32 | 0.39 | 0.413 | ≥ 3.8 | -0.23 | 0.54 | 0.678 | ≥ 3.3 | -1.17 | 0.90 | 0.193* |
| Vil. night soil sum. |  |  |  |  |  |  |  |  |  |  |  |  |
|  | ≤ 7.3 | Ref |  |  | ≤ 6.39 | Ref |  |  | ≤ 11.2 | Ref |  |  |
|  | 7.4 – 37 | 1.18 | 0.38 | 0.002* | 6.4 – 14.4 | -0.12 | 0.41 | 0.766 | 11.3 – 27.9 | 2.48 | 1.11 | 0.025* |
|  | ≥ 38 | 1.06 | 0.38 | 0.005* | ≥ 14.5 | -1.80 | 0.66 | 0.006* | ≥ 28 | 1.95 | 1.14 | 0.087* |
| Vil. night soil winter |  |  |  |  |  |  |  |  |  |  |  |  |
|  | ≤ 23.6 | Ref |  |  |  |  |  |  |  |  |  |  |
|  | 23.7 – 42.6 | -0.69 | 0.44 | 0.120* |  |  |  |  |  |  |  |  |
|  | ≥ 42.7 | 1.04 | 0.32 | 0.001* |  |  |  |  |  |  |  |  |
| HH night soil winter* |  |  |  |  |  |  |  |  |  |  |  |  |
|  | 0 | 0.90 | 0.56 | 0.105* |  |  |  |  |  |  |  |  |
|  | 1-20 | Ref |  |  |  |  |  |  |  |  |  |  |
|  | 21 – 59 | 1.11 | 0.59 | 0.060* |  |  |  |  |  |  |  |  |
|  | ≥ 60 | 0.96 | 0.58 | 0.097* |  |  |  |  |  |  |  |  |

^a^ PE = Point estimate

^b^ p.f.p = perfect failure predicted

* Between group difference (p<0.05). In the case that the tertiles did not show evidence of any moderate difference (p<0.2) between one or more groups, quartiles were tried. When still no difference was found between groups, this was noted in results Table 4.

**Figure S1. Supplemental analysis assessing changes over time.** Two additional RF model iterations were run for each collection year that only included those predictors that were available in all three of the collection years. The top ten predictors for these two iterations were given a score of 1-10, and the summed scores were used to determine the variable ranking 1^st^ (top predictor) – 10^th^ for each collection year, as well as a final variable ranking “all year score” that summed the rankings across all six iterations (two per collection year) conducted.

|  | **2007** | **2010** | **2016** | **All year score** |
| --- | --- | --- | --- | --- |
| ***Physical characteristics*** |  |  |  |  |
| Number of hatch tests |  | 2 |  | 9 |
| County of residence |  |  |  |  |
| ***Infection*** |  |  |  |  |
| Human infection: household count |  | 9 |  |  |
| Human infection: village prevalence | 5 | 1 |  | 4* |
| ***Socio-economic indicators*** |  |  |  |  |
| Village prevalence of improved sanitation |  |  | 10 |  |
| Household has improved sanitation (y/n) |  |  |  |  |
| Village mean asset score (0-9) | 10 |  | 4 | 10 |
| Household asset score (0-9) |  |  |  |  |
| ***Animal ownership*** |  |  |  |  |
| Village prevalence of cat ownership |  |  | 9 |  |
| Household cat ownership |  |  |  |  |
| Village prevalence of dog ownership |  |  | 6 |  |
| Household bovine ownership |  |  |  |  |
| Village prevalence of bovine ownership | 8 |  | 1* | 8 |
| ***Agriculture*** |  |  |  |  |
| Village mean rice area | 6 | 5 | 7* | 6 |
| Household rice area |  | 6 |  |  |
| Village mean dry summer crop area | 2 | 10 | 7* | 7 |
| Household dry summer crop area | 4 | 4 |  | 4* |
| Village mean winter crop area | 1 | 8 | 3 | 2 |
| Household winter crop area | 3 | 7 | 5 | 3 |
| Village night soil rice: mean # buckets |  |  |  |  |
| Household night soil rice: # buckets |  |  |  |  |
| Village night soil summer dry crop: mean # buckets | 7 | 3 | 1* | 1 |
| Household night soil summer dry crop: # buckets |  |  |  |  |
| Village night soil winter crop: mean # buckets |  |  |  |  |
| Household night soil winter crop: # buckets | 9 |  |  |  |
| * \| = Tied for importance rank; \| = Not ranked 1-10 for the given model/year **Color key: Variable importance rankings (1^st^-10^th^) by scale** \| \| \| \| \|; **Individual** \| 1^st^ – 2^nd^ \| 3^rd^ – 4^th^ \| 5^th^ – 6^th^ \| 7^th^ – 8^th^ \| 9^th^ – 10^th^; **Household** \| 1^st^ – 2^nd^ \| 3^rd^ – 4^th^ \| 5^th^ – 6^th^ \| 7^th^ – 8^th^ \| 9^th^ – 10^th^; **Village** \| 1^st^ – 2^nd^ \| 3^rd^ – 4^th^ \| 5^th^ – 6^th^ \| 7^th^ – 8^th^ \| 9^th^ – 10^th^ | | | | |

**Figure S2. Correlation matrix for 2007 predictors.** A correlation matrix for predictors included in the 2007 RF models is provided to highlight those predictors whose relative variable ranking positions may be less reliable due to correlation with other influential predictors. Only predictors with a correlation coefficient of < -0.499 or > 0.499 are included. The 2007 correlation matrix demonstrates that there are some strongly correlated predictors, particularly in the agricultural predictor category, that may be impacting their relative importance rankings.

|  | County | Hatch tests | HH asset | HH rice crop area | HH sum. crop area | HH win. crop area | HH NS sum. crop | Vil. asset | Vil cat own | Vil. bov. own | Vil hum. Inf. prev. | Vil rice area | Vil. sum. crop area | Vil. win. crop area | Vil. NS sum. crop | Vil. NS win. crop | Vil NS Rice crop |
| --- | --- | --- | --- | --- | --- | --- | --- | --- | --- | --- | --- | --- | --- | --- | --- | --- | --- |
| County | 1.0 |  |  |  |  |  |  |  |  |  |  |  |  |  |  |  |  |
| Hatch tests | 0.553 | 1.0 |  |  |  |  |  |  |  |  |  |  |  |  |  |  |  |
| HH asset |  |  | 1.0 |  |  |  |  |  |  |  |  |  |  |  |  |  |  |
| HH rice area |  |  |  | 1.0 |  |  |  |  |  |  |  |  |  |  |  |  |  |
| HH sum. crop area | 0.578 |  |  |  | 1.0 |  |  |  |  |  |  |  |  |  |  |  |  |
| HH win. crop area |  |  |  |  | 0.559 | 1.0 |  |  |  |  |  |  |  |  |  |  |  |
| HH NS sum. Crop |  |  |  |  | 0.594 |  | 1.0 |  |  |  |  |  |  |  |  |  |  |
| Vil Asset | -0.728 |  | 0.576 |  | -0.501 |  |  | 1.0 |  |  |  |  |  |  |  |  |  |
| Vil Cat Own |  |  |  |  |  |  |  | -0.565 | 1.0 |  |  |  |  |  |  |  |  |
| Vil Bov. Own |  |  |  |  |  |  |  | -0.520 |  | 1.0 |  |  |  |  |  |  |  |
| Vil Hum. Inf. Prev. |  |  |  |  |  |  |  |  |  |  | 1.0 |  |  |  |  |  |  |
| Vil rice area | -0.664 |  |  | 0.691 | -0.575 |  |  | 0.544 |  |  |  | 1.0 |  |  |  |  |  |
| Vil sum. crop area | 0.799 |  |  |  | -0.699 |  |  | -0.632 |  |  |  | -0.696 | 1.0 |  |  |  |  |
| Wil win. crop area |  |  |  |  |  |  |  |  |  |  |  |  | 0.648 | 1.0 |  |  |  |
| Vil NS sum. crop | 0.674 |  |  |  | 0.644 |  |  | -0.626 |  |  |  | -0.599 | 0.814 | 0.712 | 1.0 |  |  |
| Vil NS win. Crop |  |  |  |  |  |  |  |  |  |  |  |  | 0.547 |  |  | 1.0 |  |
| Vil NS rice crop |  |  |  |  |  |  |  |  |  |  | 0.598 |  |  |  |  |  | 1.0 |
| **Color Key:** Moderate positive correlation \| 0.50 – 0.599 \| Moderate negative correlation \| -0.599– -0.50; Strong positive correlation \| 0.60 – 0.799 \| Strong negative correlation \| -0.799 – -0.60; Very strong positive correlation \| ≥ 0.80 \| Very strong negative correlation \| ≤ -0.80 | | | | | | | | | | | | | | | | | |

**Figure S3. Correlation matrix for 2010 predictors.** A correlation matrix for predictors included in the 2010 RF models is provided to highlight those predictors whose relative variable ranking positions may be less reliable due to correlation with other influential predictors. Only predictors with a correlation coefficient of < -0.499 or > 0.499 are included. The 2010 correlation matrix demonstrates that there are just a few strongly correlated predictors in the agricultural predictor category. As well as the socio-economic indicator category that may be impacting relative importance rankings.

|  | County | HH rice crop area | HH sum. crop area | HH win. crop area | HH NS sum. crop | HH NS win. crop | Vil. asset | Vil cat own | Vil. bov. own | Vil pigs | Vil rice area | Vil. sum. crop area | Vil. win. crop area | Vil. NS sum. crop | Vil. NS win. crop |
| --- | --- | --- | --- | --- | --- | --- | --- | --- | --- | --- | --- | --- | --- | --- | --- |
| County | 1.0 |  |  |  |  |  |  |  |  |  |  |  |  |  |  |
| HH rice area |  | 1.0 |  |  |  |  |  |  |  |  |  |  |  |  |  |
| HH sum. crop area |  |  | 1.0 |  |  |  |  |  |  |  |  |  |  |  |  |
| HH win. crop area |  |  | 0.575 | 1.0 |  |  |  |  |  |  |  |  |  |  |  |
| HH NS sum. Crop |  |  |  |  | 1.0 |  |  |  |  |  |  |  |  |  |  |
| HH NS win. Crop |  |  |  |  | 0.565 | 1.0 |  |  |  |  |  |  |  |  |  |
| Vil Asset | -0.771 |  |  |  |  |  | 1.0 |  |  |  |  |  |  |  |  |
| Vil Cat Own | 0.589 |  |  |  |  |  | -0.533 | 1.0 |  |  |  |  |  |  |  |
| Vil Bov. Own | 0.556 |  |  |  |  |  | -0.525 |  | 1.0 |  |  |  |  |  |  |
| Vil pigs |  |  |  |  |  |  |  |  |  | 1.0 |  |  |  |  |  |
| Vil rice area |  | 0.553 |  |  |  |  |  |  |  |  | 1.0 |  |  |  |  |
| Vil sum. crop area | 0.576 |  |  |  |  |  | -0.569 | 0.535 |  |  |  | 1.0 |  |  |  |
| Vil win. crop area |  |  |  |  |  |  |  | 0.789 |  | 0.515 |  | 0.675 | 1.0 |  |  |
| Vil NS sum. crop |  |  |  |  |  |  |  |  |  |  |  | 0578 |  | 1.0 |  |
| Vil NS win. crop |  |  |  |  |  | 0.509 |  |  |  |  |  |  |  | 0.878 | 1.0 |
| **Color Key:** Moderate positive correlation \| 0.50 – 0.599 \| Moderate negative correlation \| -0.599– -0.50; Strong positive correlation \| 0.60 – 0.799 \| Strong negative correlation \| -0.799 – -0.60; Very strong positive correlation \| ≥ 0.80 \| Very strong negative correlation \| ≤ -0.80 | | | | | | | | | | | | | | | |

**Figure S4. Correlation matrix for 2016 predictors.** A correlation matrix for predictors included in the 2016 RF models is provided to highlight those predictors whose relative variable ranking positions may be less reliable due to correlation with other influential predictors. Only predictors with a correlation coefficient of < -0.499 or > 0.499 are included. The 2016 correlation matrix demonstrates that there are several strongly correlated predictors across the different predictor categories that may be impacting relative importance rankings for the 2016 RF models.

|  | County | Hatch tests | HH hum. inf. prev | HH rice crop area | HH win. crop area | HH NS sum. crop | HH NS rice crop | Vil hum inf prev | Vil. asset | Vil toilet imp | Vil. dog own | Vil cat own | Vil pigs own | Vil rice crop area | Vil. sum. crop area | Vil. win. crop area | Vil. NS sum. crop | Vil. NS win. crop | Vil. NS rice crop |
| --- | --- | --- | --- | --- | --- | --- | --- | --- | --- | --- | --- | --- | --- | --- | --- | --- | --- | --- | --- |
| County | 1.0 |  |  |  |  |  |  |  |  |  |  |  |  |  |  |  |  |  |  |
| Hatch tests | 0.615 | 1.0 |  |  |  |  |  |  |  |  |  |  |  |  |  |  |  |  |  |
| HH hum. Inf. | -0.625 |  | 1.0 |  |  |  |  |  |  |  |  |  |  |  |  |  |  |  |  |
| HH rice area |  |  |  | 1.0 |  |  |  |  |  |  |  |  |  |  |  |  |  |  |  |
| HH win. crop area |  |  |  | 0.806 | 1.0 |  |  |  |  |  |  |  |  |  |  |  |  |  |  |
| HH NS sum. crop |  |  |  |  |  | 1.0 |  |  |  |  |  |  |  |  |  |  |  |  |  |
| HH NS rice crop |  |  |  |  |  | 0.834 | 1.0 |  |  |  |  |  |  |  |  |  |  |  |  |
| Vil. hum. inf prev. | -0.839 | -0.641 | 0.661 |  |  |  |  | 1.0 |  |  |  |  |  |  |  |  |  |  |  |
| Vil asset |  |  |  |  |  |  |  |  | 1.0 |  |  |  |  |  |  |  |  |  |  |
| Vil toilet Imp. |  |  |  |  |  |  |  |  |  | 1.0 |  |  |  |  |  |  |  |  |  |
| Vil. dog own |  |  |  |  |  |  |  |  |  |  | 1.0 |  |  |  |  |  |  |  |  |
| Vil cat Own |  |  |  |  |  |  |  |  |  |  | 0.800 | 1.0 |  |  |  |  |  |  |  |
| Vil pigs | -0.502 |  | 0.611 |  |  |  |  | 0.838 | -0.563 |  |  |  | 1.0 |  |  |  |  |  |  |
| Vil rice area |  |  |  |  |  |  |  | 0.669 | -0.635 |  |  | 0.522 | 0.858 | 1.0 |  |  |  |  |  |
| Vil sum. crop area |  |  |  |  |  |  |  | 0.501 | -0.612 |  |  | 0.721 | 0.801 | 0.678 | 1.0 |  |  |  |  |
| Vil win. crop area |  |  |  |  |  |  |  | 0.590 | -0.653 |  |  | 0.640 | 0.820 | 0.955 | 0.774 | 1.0 |  |  |  |
| Vil NS sum. crop |  |  |  |  |  |  |  |  |  | -0.645 |  |  |  |  |  |  | 1.0 |  |  |
| Vil NS win. crop | 0.649 |  |  |  |  |  |  | -0.546 |  | -0.814 |  |  |  |  |  |  | 0.578 | 1.0 |  |
| Vil. NS rice crop |  |  |  |  |  |  |  |  |  |  |  |  |  |  |  |  | 0.581 |  | 1.0 |
| **Color Key:** Moderate positive correlation \| 0.50 – 0.599 \| Moderate negative correlation \| -0.599– -0.50; Strong positive correlation \| 0.60 – 0.799 \| Strong negative correlation \| -0.799 – -0.60; Very strong positive correlation \| ≥ 0.80 \| Very strong negative correlation \| ≤ -0.80 | | | | | | | | | | | | | | | | | | | |
